# Severity assessment of single dose Oxford–AstraZeneca vaccinated individuals infected with SARS CoV-2 in the Southeast Bangladesh

**DOI:** 10.1101/2021.05.10.21256529

**Authors:** Eaftekhar Ahmed Rana, Pronesh Dutta, Md. Sirazul Islam, Tanvir Ahmad Nizami, Tridip Das, Sharmin Chowdhury, Goutam Buddha Das

**Author notes:** **Corresponding author:** Md. Sirazul Islam. **Co-authors:** EAR, PD, TAN, TD, SC, GBS.

## Abstract

The present global endeavor to uncover the most effective vaccines against severe acute respiratory syndrome coronavirus (SARS-CoV-2) that can tremendously prevent transmission, infection and significantly reduce public health risk. COVID-19 vaccination program is underway in different parts of the world including Bangladesh but till to date there is no available health data revealed among the vaccinated peoples. We conducted a cross-sectional study from February 15 to April 15, 2021 to assess the health status of 1^st^ dose Oxford-AstraZeneca vaccinated individuals infected with SARS CoV-2. Standard virological method, real-time reverse transcriptase-polymerase chain reaction (RT-qPCR) was performed to detect SARS-CoV-2 and the different health parameters from vaccinated individuals were collected through direct mobile phone contact using pre-structured questionnaires. A total of 6146 suspected samples were tested and 1752 were found positive for SARS-CoV-2, of them 200 individuals were identified who received 1^st^ dose of COVID-19 vaccine. Within the test period, majority of male (65.6%) and female (34.4%) carried moderate numbers of viruses which comprise between 30.01-35 cyclic threshold (ct) values. Among the vaccinated individuals, 165 (82.5%; 95% CI: 76.51 - 87.5) persons were not hospitalized and 177 (88.5%; 95% CI: 83.24 - 92.57) did not show any respiratory difficulties. Only a few (16) (8%; 95% CI: 4.64 - 12.67) of COVID-19 positive patients needed extra oxygen support and 199 (99.5%; 95% CI: 97.25 - 99.99) individuals didn’t require any intensive care unit (ICU) interference. Overall, oxygen saturation was recorded around 96.8% and respiratory difficulties did not extend more than 5 days, irrespective of age and sex during the infection period. Within the vaccinated COVID-19 positive individuals 113 (56.5%; 95% CI: 49.33 - 63.48) and 111(55.5%; 95% CI: 48.32 - 62.51) persons have normal physiological taste and smell. However, we have found a larger proportion of vaccinated persons (129) (64.5%; 95% CI: 57.44 - 71.12) carrying different comorbidity, among them high blood pressure 36 (27.9%; 95% CI, 20.37 - 36.48) and diabetes 32 (24.8%; 95% CI: 17.63 - 33.18) were found more prevalent. Moreover, the significant finding of the present study was 199 (99.5%; 95% CI: 97.25 - 99.99) vaccinated individuals survived with good health conditions and became negative in RT-qPCR. The authors suggest that health risk assessment among the COVID-19 vaccinated persons when infected with SARS-CoV-2 is crucial and time demanding task for the whole world. However, the present study illustrates that the administration of the 1st dose Oxford-AstraZeneca vaccine significantly reduces health risk during the COVID-19 infection period.

## Introduction

Severe acute respiratory syndrome coronavirus (SARS-CoV-2) pandemic is envisaged as number one global public health crisis due to its high morbidity and drastic fatality rate. Since the reporting of the outbreak, it has infected over 153 million of which over 3.2 million people died (Worldometer 2021). Longer survival rate of virus in different environmental conditions and unprecedented speed of transmission from human-to-human can aggravate the present ongoing outbreak situations (Negahdaripour, 2020). Additionally, the virus causes severe flu-like symptoms with greater respiratory difficulties and reports that one infected or carrier individual can easily infect others (J. Xu et al., 2020). However, the estimated reproductive number (R_0_) of SARS-CoV-2 is 2.2, i.e., one COVID-19 individual is able to transmit the virus to 2.2 other healthy individuals (Li Q et al., 2020). To overcome the devastating effects of the virus on human health, a highly effective vaccine is a crying need. For COVID-19 prevention different vaccine platforms such as nucleic acid vaccines, recombinant protein vaccines, viral vector-based vaccines and whole virus vaccines are targeted (Chen et al., 2020). To date, 180 different vaccine candidates are currently developing vaccines against SARS-CoV-2 (Krammer, 2020) of which 58 vaccines have been developed by different institutes and companies. Among them some vaccines are under clinical trials (Knoll and Wonodi, 2021) and few of them got permission for mass vaccination by the World Health Organization (WHO) for the successful COVAX programs co-leaded by Gavi, CEPI and WHO. Oxford-AstraZeneca chimpanzee adenovirus vectored vaccine (ChAdOx1 nCoV-19) received their license for vaccination program with a reported 90% efficacy against SARS CoV-2 after a second dose (Knoll and Wonodi, 2021). Nowadays, immunization campaign is continued in several countries including Bangladesh (DGHS 2021) irrespective of age and sex although senior citizens are experiencing a priority. The Bangladesh government started a free vaccination campaign over the country against COVID-19 using the Oxford-AstraZeneca vaccine received from the Serum Institute of India. In Chattogram division the commencement was from February 7, 2021 (The Business Standard 2021). However, the magnitude of vaccine response to the virus particle is widely varied. The significant consideration is the re-infection of previously infected or vaccinated individuals with the same virus is possible due to its high and rapid mutation rate as well as the nature of viruses (Hansen et al., 2021). However, human coronavirus does not induce lifelong immunity and antibody response due to rapid fall of humoral immunity (Amanat and Florian Krammer,2020). Moreover, the measurement of protective antibody titer against SARS-CoV-2 after vaccination is still underdeveloped. Unfortunately, after receiving the first dose (approximately 5 × 10^10^ viral particles) of the Oxford-AstraZeneca vaccine, a number of vaccinated people were re-infected with SARS-CoV-2. Considering the present COVID-19 pandemic crisis and vaccination status, we aimed to find out the percentages of first dose vaccinated individuals re-infected with SARS-CoV-2 and assess their health risk during the infection period.

## Materials and Methods

### Ethical approval and authorization

All of the samples were collected as a part of COVID-19 diagnosis that was provided by every individual with their own interest and consent. However, before the sample collection minimum discomfort was maintained in every patient and verbal permission was taken before the collection of COVID-19 vaccination history as well as health related information during the infection period. Finally, authorization from the Research and Extension director of Chattogram Veterinary and Animal Sciences University (CVASU) was taken to conduct the present study.

### Study area

The present study was conveyed in the greater Chattogram division of Bangladesh which comprises 11 districts (BNP 2021/13). Geographically, it is located in the southeast part of Bangladesh and well recognized as one of the major seaports of the country (Rana et al., 2020). However, among the 11 districts, only four districts namely Khagrachhari, Rangamati, Bandarban and Chattogram were included in our study.

### Study Population and period

Any individual received first dose of COVID-19 vaccine from any health care center in Chattogram division irrespective of age and sex from the study area was included in the current study. The study was conducted during the campaign of 1st dose vaccination which began from February 15, 2021 to April 15, 2021 (2 months) before the starting of COVID-19 2nd dose vaccination campaign.

### Sample Collection

Nasal and oropharyngeal samples of the suspected individuals within the study area were sent to COVID-19 detection laboratory of Chattogram Veterinary and Animal Sciences University (CVASU) through Chattogram Medical College (CMC) and Bangladesh Institute of Tropical and Infectious Diseases (BITID). Individual samples were collected in separate collection tubes containing viral transport media (VTM) maintaining the WHO guidelines (WHO, 2020). Samples were preserved into −80^0^C temperature immediately after collection and sent to the COVID-19 detection laboratory maintaining the proper cool chain.

### Molecular Diagnosis

After receiving the suspected samples from authorities, individual samples were tested for detection of SARS-CoV-2 by RT-qPCR method. Viral RNA was extracted by using sample release reagent (SanSure Biotech, Ref. No - S1014E), following the manufacturer’s indications. Novel Coronavirus (2019-nCoV) nucleic acid diagnostic kit (PCR-Fluorescence Probing, Ref. No-S3102E) (Sansure Biotech 2019) was used to detect the N gene (ROX channel) and ORF1ab region (FAM channel) of SARS-CoV-2 from extracted samples’ RNA. To regulate the PCR inhibition, human RNA targeting *P* gene (CY 5 channel) was used as an internal control. RT-qPCR was performed on a QuantStudio™ 5 PCR system with the version 1.5.1 for analysis. Any samples showed ≤ 40cyclic threshold (ct) value which confirmed as positive for COVID-19.

### Data Collection

After laboratory confirmation of the SARS-CoV-2 positive cases, we traced each COVID-19 patient over the phone. Only 1st dose vaccinated COVID-19 positive individuals were included to collect data on vaccination history and health related demographic information during the infection period through a structured questionnaire. Vaccinated COVID-19 positive patients were traced until becoming free from viral infection as well as post COVID-19 complication and, COVID-19 negative test results. All data was sorted and coded in Microsoft Excel 2016^®^ excel sheet for further summary and analysis.

### Statistical Analysis

After sorting, all the data were inserted in STATA-IC 13^®^ software to perform statistical analysis. Descriptive analysis was performed to calculate the prevalence of target outcome. The prevalence of SARS-CoV-2 was calculated considering the number of COVID-19 positive cases as the numerator divided by the total number samples as the denominator. The 95% confidence interval of the prevalence values was calculated by the modified Wald method using the Graph Pad Quickcalcs Online tool (www.graphpad.com/quickcalcs/).

## Results

### Prevalence of COVID-19 vaccinated patients

A total of 6146 suspected samples were tested by targeting SARS-CoV-2 virus within the study period, among them 1752 (28.51%; 95% CI: 27.38 - 29.65) samples were found positive for COVID-19. From the positive cases, we found 200 (11.42%; 95% CI: 9.96 - 13) individuals received the 1^st^ dose of Oxford-AstraZeneca vaccine. Within the vaccinated COVID-19 positive individuals 134 (67%; 95% CI: 60.02 - 73.47) were found male and the remaining 66 (33%; 95% CI: 26.53 - 39.98) were female.

In our study, we observed 165 (82.5%; 95% CI: 76.51-87.5), 1^st^ dose vaccinated COVID-19 positive patients were not admitted to hospital where according to sex, 110 (82.09%; 95% CI: 74.53 - 88.17) male and 55 (83.33%; 95% CI:72.13-91.37) female took treatment within home.

### Prevalence of COVID-19 vaccinated patients in different ct value categories

All of the vaccinated SARS CoV-2 infected patients were categorized according to viral load which is based on cyclic threshold (ct) value of tested RT-qPCR **(Figure 1)**. Among the 200 patients, 18 (9%; 95% CI: 5.42 - 13.85) had exhibit high viral loads irrespective of age and sex which comprise below or equal 20 ct value and occupied category 1, where 20.01 - 25 ct value, 25.01 - 30 ct value, 30.01 - 35 ct value and 35.01 - 40 ct value were in category 2, category 3, category 4 and category 5 accordingly. And most of the patients, 61 (30.5%; 95% CI: 24.2 - 37.39) were found in category 4. Within the age ranges, 40 - 49 years old individuals were found to carry high viral load during the test period.

**Figure 1:**
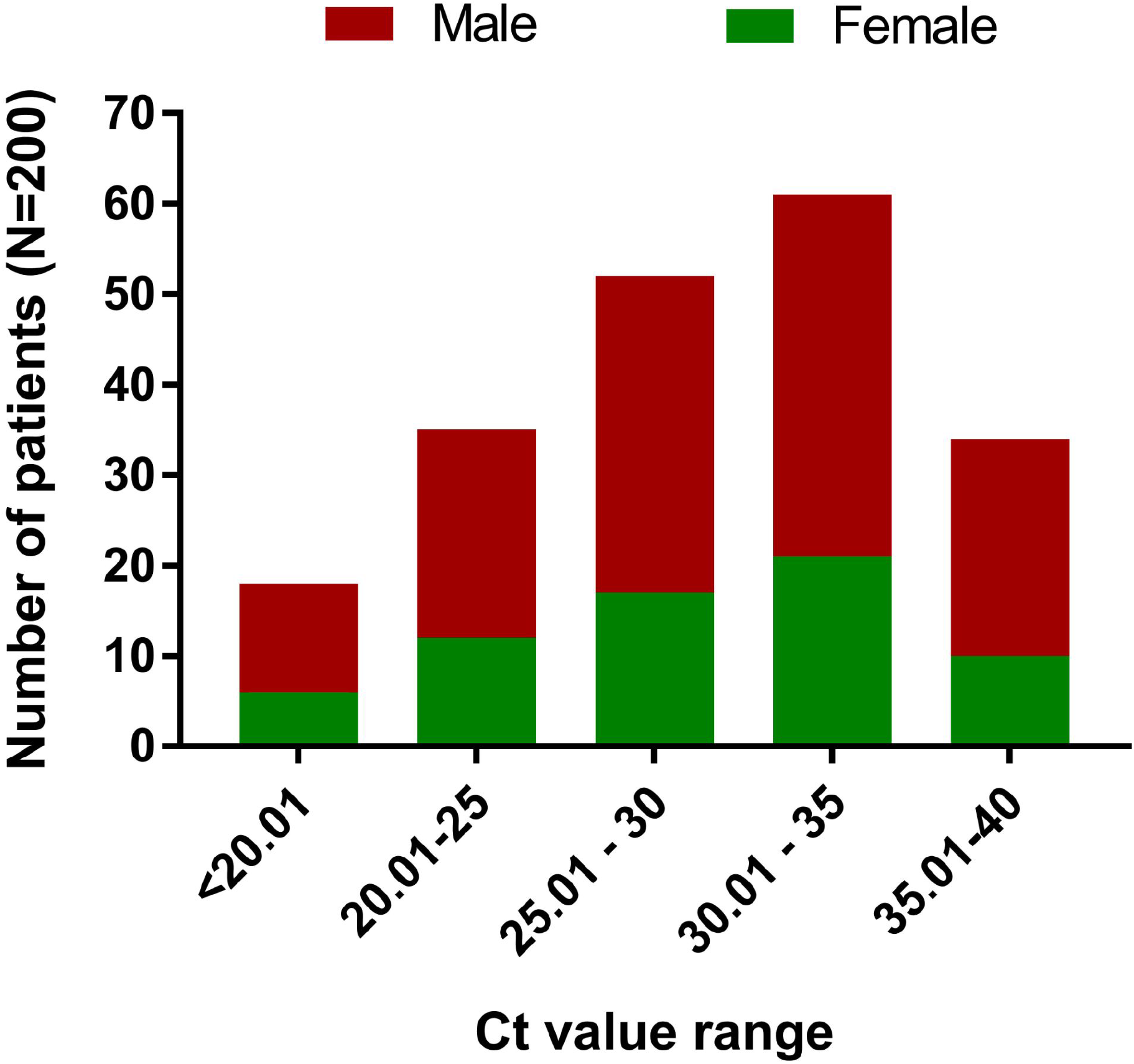
Category of SARS CoV-2 infected vaccinated patients based on viral load using cyclic threshold (ct) value of RT-qPCR. Where category 1: ≤20 (ct); category 2: 20.01-25 (ct); category 3: 25.01-30 (ct); category 4: 30.01-35 (ct) and category 5: 35.01-40 (ct).

### Prevalence of general physiological symptoms with parameters

The primary and well defined symptoms of the coronavirus are likely fever, coughing and sneezing. In our findings, we noticed 144 (72%; 95% CI: 65.23 - 78.1) individuals appear fever with variable ranges while 182 (91%; 95% CI: 86.15-94.58) and 89 (44.5%; 95% CI: 37.49-51.68) COVID 19 patients didn’t have any sneezing and coughing during the infection period **(Table 1)**. Irrespective of ages and sex, it was observed that within the vaccinated individulas the common symptoms of sneezing and coughing were not extended more than 3 days and 7 days, respectively. However, 113 (56.5%; 95% CI: 49.33 - 63.48) and 111 (55.5%; 95% CI: 48.32 - 62.51) vaccinated individuals had their normal physiological taste and smell function during the infection period.

**Table 1:**
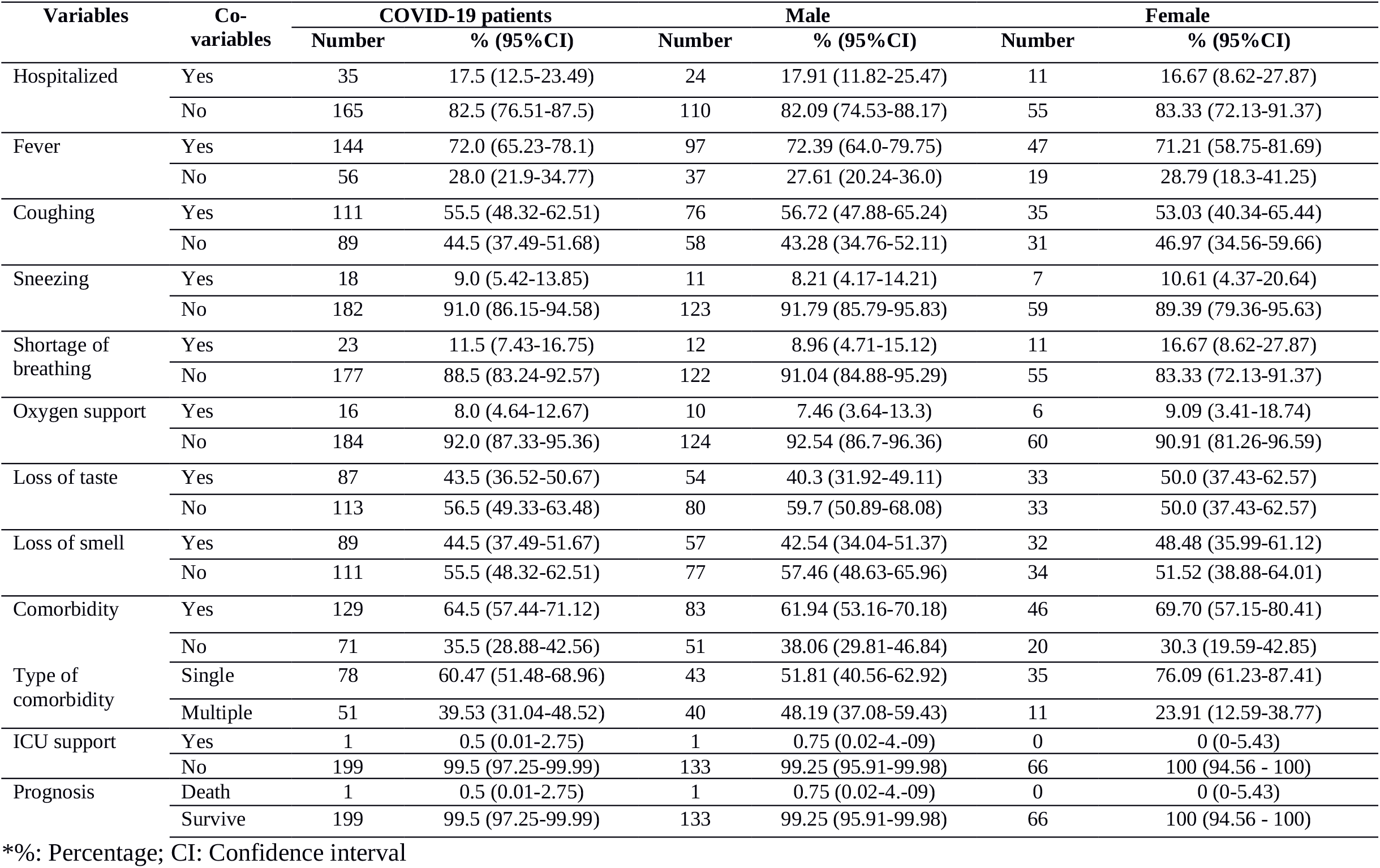
Prevalence of different physiological conditions of vaccinated COVID-19 individuals during SARS CoV-2 infection period

### Respiratory difficulties and oxygen saturation

Shortness of breathing is one of the most significant symptoms of the COVID-19 patients. We found 177 (88.5%; 95% CI: 83.24 - 92.57) vaccinated patients didn’t express any breathing difficulties, of them 122 (91.04%; 95% CI: 84.88 - 95.29) and 55 (83.3%; 95% CI: 72.13 - 91.3) male and female, respectively found free from dyspnea. Moreover, in general breathing difficulties of SARS-CoV-2 infected patients were persistent around 5 days **(Table 2)**. Interestingly, 184 (92%; 95% CI: 87.33-95.36) COVID-19 positive vaccinated patients didn’t require any extra oxygen support from our source. The overall oxygen saturation levels of vaccinated COVID-19 patients were found 96.8% (95% CI: 96.5-97.2), where it was 97% (95% CI: 96.5-97.4) in male and 96.5% (95% CI: 95.9-97.1) in females.

**Table 2:** Different physiological parameters of vaccinated COVID-19 patients during infection period.

| Variables | COVID-19 patients |  | Male |  | Female |  |
| --- | --- | --- | --- | --- | --- | --- |
| | Mean ± SE (95% CI) | Min - Max | Mean ± SE (95% CI) | Min - Max | Mean ± SE (95% CI) | Min- Max |
| COVID-19 positive result after onset of vaccination (days) | 32 ± 1.2 (29.7-34.3) | 1 - 61 | 32 ± 1.4 (29.1-34.8) | 1 - 61 | 32 ± 2 (28.1-35.9) | 3 - 57 |
| Duration of fever (days) | 3.9 ± 0.2 (3.5-4.4) | 1-15 | 3.9 ± 0.3 (3.4-4.5) | 1 - 10 | 3.9 ± 0.4 (3.1-4.6) | 1 - 15 |
| Body temperature ($^{\circ}$ Fahrenheit) | 100.9 ± 0.1 (100.7-101.2) | 99 - 104 | 100.9 ± 0.1 (100.6-101.2) | 99 - 104 | 101 ± 0.2 (100.7-101.3) | 99 - 104 |
| Duration of coughing (days) | 6.4 ± 0.4 (5.5-7.2) | 1 - 25 | 6.5 ± 0.6 (5.4-7.6) | 1 - 25 | 6.1 ± 0.7 (4.7-7.5) | 2 - 21 |
| Duration of sneezing (days) | 2.9 ± 0.5 (1.8-4) | 1 - 8 | 2.5 ± 0.6 (1.2-3.9) | 1 - 8 | 3.7 ± 1 (1.1-6.2) | 2 - 8 |
| Duration of shortness of breathing (days) | 4.8 ± 0.7 (3.4-6.3) | 1 - 12 | 5 ± 1.1 (2.5-7.5) | 3 - 12 | 4.6 ± 0.9 (2.4-6.8) | 1 - 8 |
| Oxygen saturation level (%) | 96.8 ± 0.2 (96.5-97.2) | 90 - 99 | 97 ± 0.2 (96.5-97.4) | 90 - 99 | 96.5 ± 0.3 (95.9-97.1) | 90 - 99 |

\*SE: Standard Error; Min: Minimum; Max: Maximum; %: Percentage; CI: Confidence interval

### Comorbidity

Within the vaccinated COVID-19 patients, a total of 129 (64.5%; 95% CI: 57.44 - 71.12) individuals carried different types of comorbidity, where hypertension (36), and diabetes (32) are found more prevalent. Among the co-morbidity patients, 51 (39.5%; 95% CI: 31.04 - 48.52) individuals were identified they carried more than one co-morbidity **(Figure 2)**. Moreover, study revealed that a significant number of male 83 (61.94%; 95% CI: 53.16 - 70.18) suffered from different types of co-morbidity than female 46 (69.7%; 95% CI: 57.15 - 80.41). In our study, we found only 1 (0.5%; 95% CI: 0.01 - 2.75) individuals died after taking the 1st dose of Oxford-AstraZeneca vaccine within the infection period.

**Figure 2:**
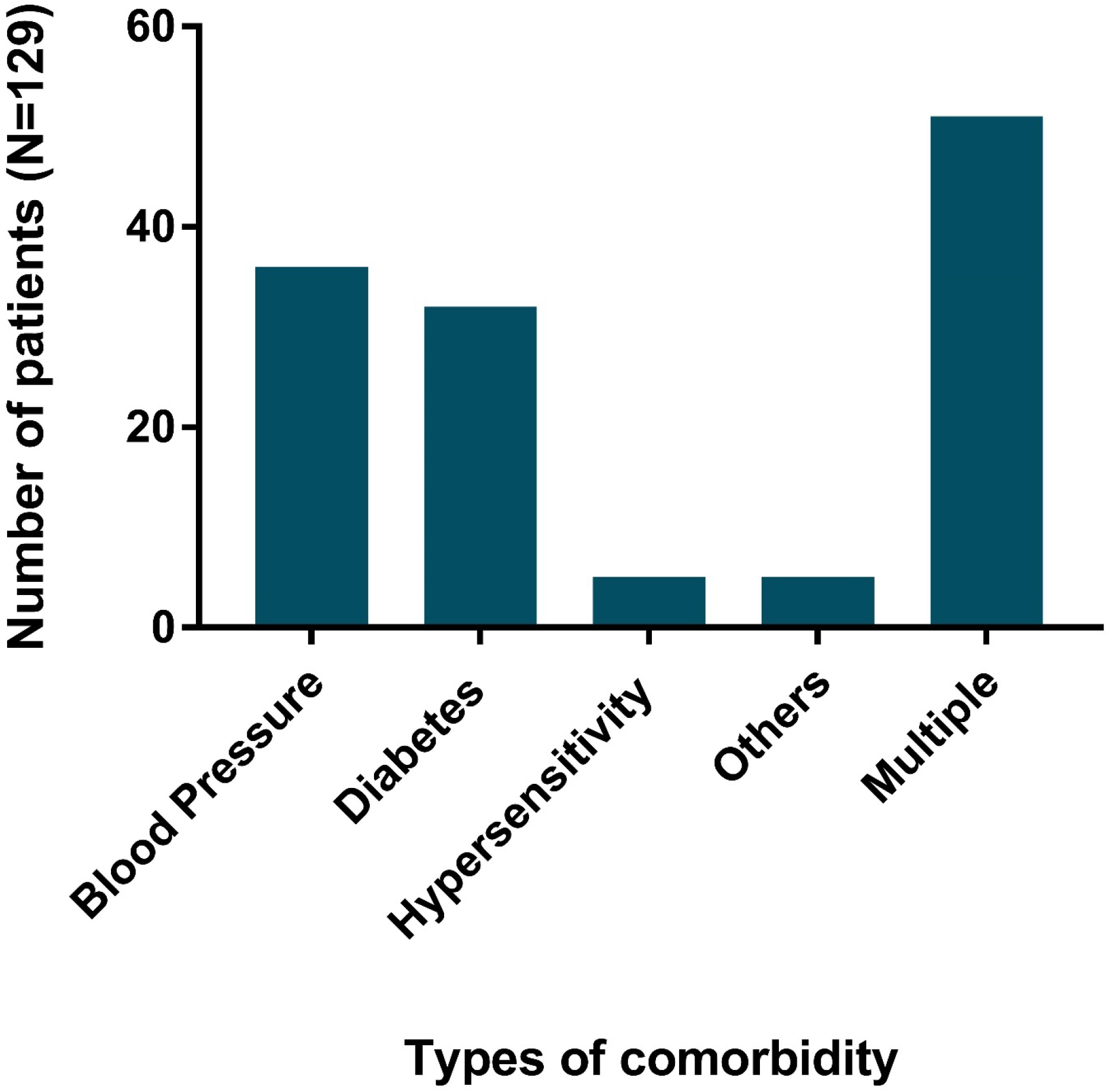
Prevalence of different comorbidity retaining COVID-19 patients infected with SARS CoV-2 after vaccination. Here, comorbidity type others include cardiac disorder, kidney disease and liver disease.

## Discussion

Assessment of the safety and efficacy of COVID-19 vaccines against the SARS-CoV-2 among the different populations is essential for an effective global pandemic response. The present study reveals the severity of single dose Oxford-AstraZeneca vaccinated people infected with SARS CoV-2 in the Southeast part of Bangladesh. The overall prevalence of positive COVID-19 individuals was 28.51% which is almost similar to 29.76% prevalence, reported in early outbreak in the same study region (Rana et al., 2020). The prevalence of 1st dose vaccinated (Oxford-AstraZeneca) individuals from the positive cases were 11.42% of which 67% and 23% were found male and female, respectively. However, infection after 1^st^ dose vaccination was also reported among health care workers in California, USA which was 2.59% at different time intervals (Keehner et al., 2021). The reason for reinfection after vaccination might be due to the frequent mutation of SARS-CoV-2 which has less protection against certain variants like the UK variant (B.1.1.7), South African variant (B.1.351), and Brazilan variant (P1/P2) which were recently detected in Bangladesh (icddr,b, 2021). Since the Oxford-AstraZeneca vaccine was designed by targeting the spike protein gene but frequent mutation of this region may alter the immunological response and fail to give protection. The newly emerged variants are said to have greater transmissibility and continuously harbour new genetic changes, which may impact on clinical manifestation and vaccine effectiveness (icddr,b, 2021). Mahdi et al, 2021 also reported two-dose regimen of ChAdOx1-nCoV19 did not show protection against mild-moderate Covid-19 caused by B.1.351 variant. However, experiments reveal that a single standard dose of Oxford AstraZeneca vaccine provided around 76% protection against symptomatic patients with COVID-19 (Wise, 2021).

Among the vaccinated positive individuals 82.5% did not need any hospital care and they received nursing staying at home. This finding supports Iacobucci, 2021 who also reported 80% reduction of hospital admission after 1^st^ dose vaccination with Oxford-AstraZeneca. The reason behind lower hospital admission rates might be due to the protection and efficacy given by the vaccine against severe clinical symptoms (Knoll and Wonodi, 2021). Body immunity developed in response to vaccines which also works to reduce the severity of infections and subside the systemic clinical manifestation thus ultimately prevent mortality (Ramasamy et al., 2020).

The present findings reported 28%, 44.5% and 91% vaccinated individuals did not show any symptoms of fever, coughing and sneezing, respectively during the infection period. The duration and severity of all symptoms were also found low. Furthermore, 56.5% and 55.5% individuals had no changes in their normal taste and smell sensation. The reason for milder symptoms of COVID-19 positive vaccinated individuals might be due to the quick immune response generated by ChAdOx1-nCoV19 maintaining a certain antibody titer that inhibit viral replication and reducing viral loads (Hung et al., 2021; Wise, 2021).

Breathing difficulties and low oxygen saturation are commonly noticed in severe COVID-19 patients. However, in our study 88.5% 1st dose vaccinated individuals did not show any sign of dyspnoea and their average oxygen saturation level was found normal (96.8%). This is because SARS-CoV-2 infection is generally mild in the majority of individuals. However, it is well defined the Oxford-AstraZeneca vaccine is developed based on SARS-CoV-2 spike protein gene which replicates inside the host cell after immunization and produces significant T-cell responses against it which prevents SARS-CoV-2 spike protein binding to angiotensin-converting enzyme 2 (ACE-2) receptor of lungs and also capable to neutralize the virus inside the host body (Ramasamy et al., 2020; Bertoletti et al, 2021). Very few vaccinated individuals develop respiratory difficulties which might be due to presence of comorbidities, secondary bacterial infection and an initial defect in antiviral host defense mechanisms (Netea et al., 2020). Recent emergence of the UK, B.1.1.7 (also called 501Y.V1) includes eight amino acid changes within the spike. One of these, N501Y (Asn 501 Tyr), increases the affinity of spike to binding its cellular target ACE-2 receptor and causes severe lung damage during the replication process. And thus underlying causes significantly reduce oxygen consumption as well as blood oxygen saturation level and probably this is the main trigger for breathing difficulties in COVID-19 positive individuals (Altmann et al., 2021; Bertoletti et al, 2021).

Presence of comorbidities are linked to severity of COVID-19 and substantially associated with significant morbidity and mortality (Ejaz et al., 2020). Although 17.5 percent vaccinated yet infected patients were admitted to the hospital, no serious health risk was observed despite presence of co-morbidities in 64.5% individuals. Among different kinds of comorbidities hypertension was found to be highest (27.9%) followed by diabetes (24.8%). SARS-CoV-2 utilizes ACE-2 receptors expressed at the surface of the host cells to access inside the cell. Certain comorbidities are associated with a potent ACE-2 receptor expression and higher release of pro-protein convertase that enhances the viral entry into the host cells (Ejaz et al., 2020). However, the presence of comorbidities especially diabetes increases susceptibility of SARS CoV-2 infection (Erener, 2020) and significantly reduces the body immunity function. Moreover, it also enhances the acute cytokine storm, pulmonary dysfunction and hypercoagulation of SARS CoV-2 infected patients (Erener, 2020). About 199 (99.5%) individuals were found alive upto negative COVID-19 test results, while only one individual with a history of kidney transplantation and presence of multiple comorbidities died.

Minimum adverse events or deaths in ChAdOx1 nCoV-19 single dose vaccine recipients were also reported by Knoll and Wonodi, 2021. Oxford AstraZeneca vaccines are found effective in reducing COVID-19 infections and protecting against severe disease in adults (Iacobucci, 2021).

However, only vaccinated individuals were considered for this study where a comparison study between vaccinated and non-vaccinated individuals is also important for an effective vaccine efficacy study. It is also better to sequence the viruses that infect the vaccinated individuals and help to identify the strain and nature as well as molecular dynamics of SARS CoV-2. The study was conducted in a certain geographical location of Bangladesh. However, elaborate studies including large number vaccinated individuals in all the divisions of Bangladesh are recommended for future studies that make clear understanding about vaccine efficacy against COVID-19.

## Conclusion

The present investigation evidently focused the SARS-CoV-2 onfall even though reception of 1^st^ dose Oxford-AstraZeneca COVID-19 vaccine. However, the hearty finding is 82.5% single dose vaccinated COVID-19 patients did not require any hospital care and 88.5% individuals did not face any respiratory difficulties. Despite the presence of different comorbidities in the vaccinated patients, only 8% of the study population required extra oxygen support and negligible (0.5%) number of patients admitted into ICU. The study concludes that vaccination with a single dose Oxford AstraZeneca vaccine significantly reduced severity and mortality of COVID-19 patients in southeast Bangladesh. Finally, we propose the studies of SARS-CoV-2 infection after vaccination should be integrated into the development of highly effective and durable immune responses producing vaccines that curb the COVID-19 pandemic.

## Data Availability

The data availabilty link is not given.

## Author’s Contribution

EAR planned, designed the study and prepared the initial draft of the manuscript. EAR, PD, MSI, TAN, and TD executed laboratory tasks, data entry and manuscript preparation. PD and TD analysed and summarised the data. SC and GBD supervised the study and revised the final manuscript. All authors read and approved the final manuscript.

## Acknowledgement

The authors acknowledge CMC and BITID for providing samples from different regions of the Chattogram division. The authors also acknowledge Fahad Hossain Palash and Md. Jahid Hasan for collecting patients’ data. The laboratory reagent facilities were supported by the Directorate General of Health Services, People’s Republic of Bangladesh. The authors sincerely acknowledge the Director, Poultry Research and Training Centre (PRTC), CVASU for laboratory logistic support. The study was funded by the Director of Research & Extension,CVASU.

